# GEO Data Sets Analysis On Mechanism of Action of IFNβ-1a Treatment in Multiple Sclerosis

**DOI:** 10.1101/2023.02.25.23286450

**Authors:** Adam Ho

## Abstract

Multiple Sclerosis (MS) is an autoimmune disease that affects millions of people worldwide and causes symptoms such as dysarthria, ataxia, and nystagmus. MS is known to be characterized by an autoimmune attack by the immune system on the myelin sheath of neurons, causing inflammation and scarring (sclerosis). In the status quo, MS is treated or alleviated by disease-modifying therapies, including beta interferons (IFNβ) and monoclonal antibodies. Yet, the mechanism of action (MOA) of IFNβ is not fully understood, and only a limited proportion of patients respond to IFNβ treatment. Mononuclear cells from therapy-naïve MS patients, IFN-β-1a-treated MS patients after 12 months from three databases on GEO are analysed to examine RNA changes that characterize both the disease and its treatment. 28 differentially expressed genes (DEGs) are identified in all three of the databases and passed the cut-off criteria. Using the 28 DEGs, we performed DAVID and PANTHER analysis, revealing that the biological process “immune response”, “defence against virus”, and “regulation of viral genome replication” are enriched. A protein interaction network for the DEGs was constructed and a protein module was identified and analysed with PANTHER, revealing “interleukin-27-mediated signalling pathway”, “regulation of ribonuclease activity”, “regulation of type III interferon production”, “cellular response to exogenous double-stranded RNA (dsRNA)”, and “ISG15-protein conjugation are enriched for >100 folds. Cytoscape analysis further identified the hub genes IFI44L, IFI44, and STAT1 and they may be important mediators in the therapeutic effect of IFNβ treatment and warrant further study. Overall, the findings of the present study provide insights into the MOA of IFNβ-1a and provide greater confidence on which genes are differentially expressed in MS before and after IFNβ-1a treatment. The results also are additional evidence for the role of viral infection in MS, a topic that is gaining interest in the MS research community.

## Introduction

MS is a chronic autoimmune disease that affects the central nervous system. It is a progressive disease characterized by inflammation, demyelination, and neurodegeneration. MS affects millions of people worldwide and has no known cure [1]. However, there are treatments that can help to slow the progression of the disease, and one of the most commonly used treatments is IFNβ [2].

The initial interest in IFNβ as a potential therapeutic option in MS was motivated primarily by the known antiviral activities of IFNβ. Subsequently, the immunomodulatory and antiproliferative properties of IFNβ were discovered [3]. Generally, IFNβ works by regulating the immune system and preventing it from attacking the myelin sheath that surrounds nerve cells. It has been shown to reduce the frequency and severity of relapses in people with MS. It can also slow the progression of disability in people with relapsing-remitting MS [1]. Mechanisms proposed on how this is achieved include inhibition of T-cell activation and proliferation, apoptosis of autoreactive T cells, induction of regulatory T cells, inhibition of leukocyte migration across the blood-brain barrier, and potential antiviral activity [4].

While IFNβ can be an effective treatment for MS, there are challenges in using it. Firstly, the ways in which IFNβ produces its therapeutic effects in MS are not yet fully understood, however, IFNβ beneficial effects are most likely associated with its immunomodulatory properties [5]. Secondly, not all people with MS respond to the treatment. It has been estimated that up to 50% of MS patients do not respond to IFN-β treatment even with regular injections of IFNβ [6]. The present study seeks to elucidate the MAO of IFNβ, specifically IFNβ-1a, in MS which would allow further stratification of patient responders.

## Method

### Data Selection

The Gene Expression Omnibus (GEO) is an online NCBI repository containing public gene expression data from a variety of studies [7-8]. GEO was searched for datasets matching “Multiple Sclerosis”, “IFNβ-1a”, and “*Homo sapiens*”. GSE26104, GSE5574, and GSE53716 are selected for this study [9-11]. In total, 20 post-treatment samples (patients who have taken IFNβ-1a up to 12 months) and 20 pre-treatment samples (before taking IFNβ-1a) are present across the three studies. However, after removing outliers, we have a total of 18 post-treatment samples and 18 pre-treatment samples (Table 1). All three studies are profiling gene expression from RNA extracted from patient blood.

**Table 1.**
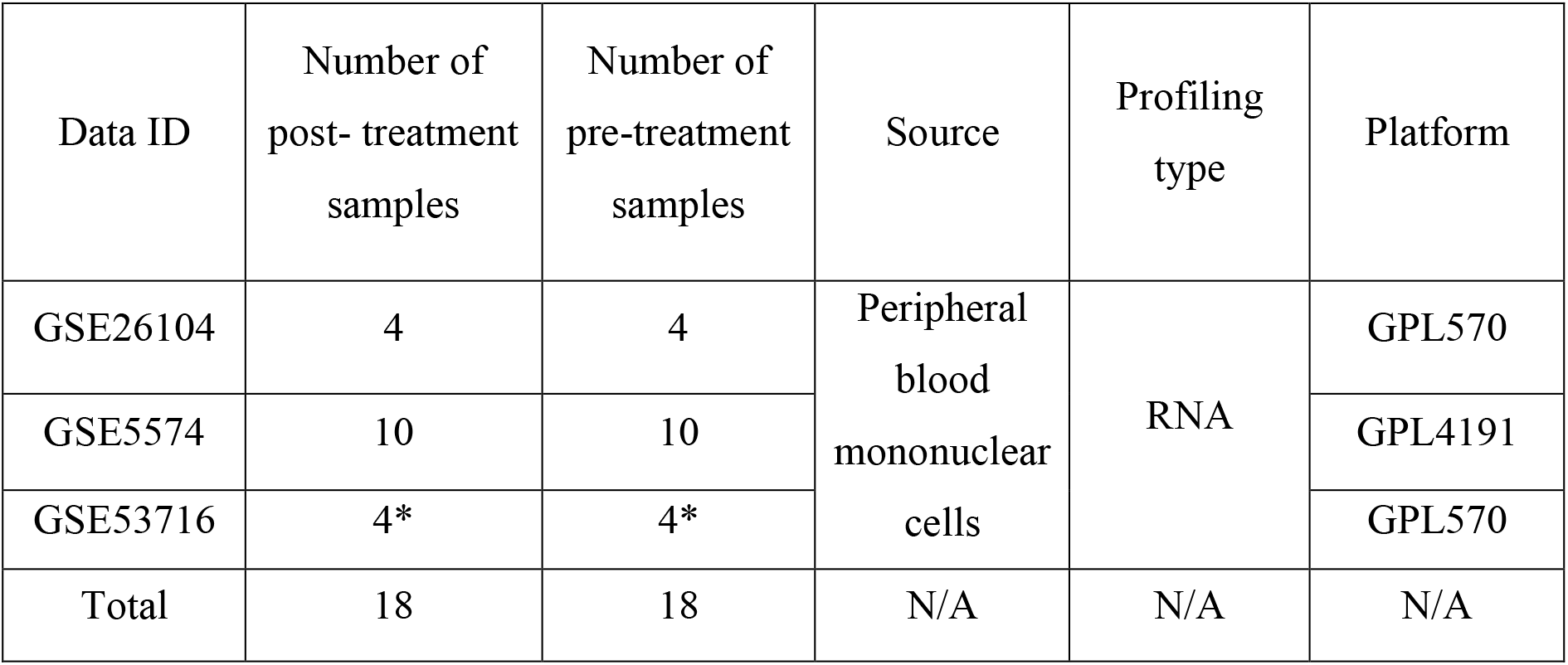
Information on GSE26104, GSE5574, and GSE53716, including the number of pre-treatment and post-treatment samples, sources, and profiling type. *Outliers are being removed in GSE52716 to ensure data normalization.

| Data ID | Number of post- treatment samples | Number of pre-treatment samples | Source | Profiling type | Platform |
| --- | --- | --- | --- | --- | --- |
| GSE26104 | 4 | 4 | Peripheral blood mononuclear cells | RNA | GPL570 |
| GSE5574 | 10 | 10 |  |  | GPL4191 |
| GSE53716 | 4* | 4* |  |  | GPL570 |
| Total | 18 | 18 | N/A | N/A | N/A |

### Differential Expression Analysis

The three datasets were analysed using GEO2R (https://www.ncbi.nlm.nih.gov/geo/geo2r/ accessed on 25 February 2023) which is provided by the Gene Expression Omnibus. GEO2R is a browser-based software that processes gene expression values and outputs a table of DEGs between two user-defined groups (pre-treatment and post-treatment, in this case) [12]. In total, several thousand genes are deemed statistically significant by GEO2R for each dataset. T-tests are used to determine p-values. GEO2R is also used to calculate fold change (FC) for each gene (in this context, of treatment expression values versus pre-treatment values). The FC is a ratio of the average expression value of a gene in one group divided by the average expression value in a different group.

GEO2R was also used to verify a normal distribution of gene expression values. The samples from GSE2614 and GSE5574 suggest excellent performance of normalization since the median of the expression value was almost on the same line (Figure 1a). However, 2 samples are removed from GSE53716 pre-sample and post-treatment samples respectively to ensure data normalization (Figure 1b). Normalization is essential to account for factors that affect the number of reads mapped to a gene, like length, GC-content, and sequencing depth [13].

**Figure 1.**
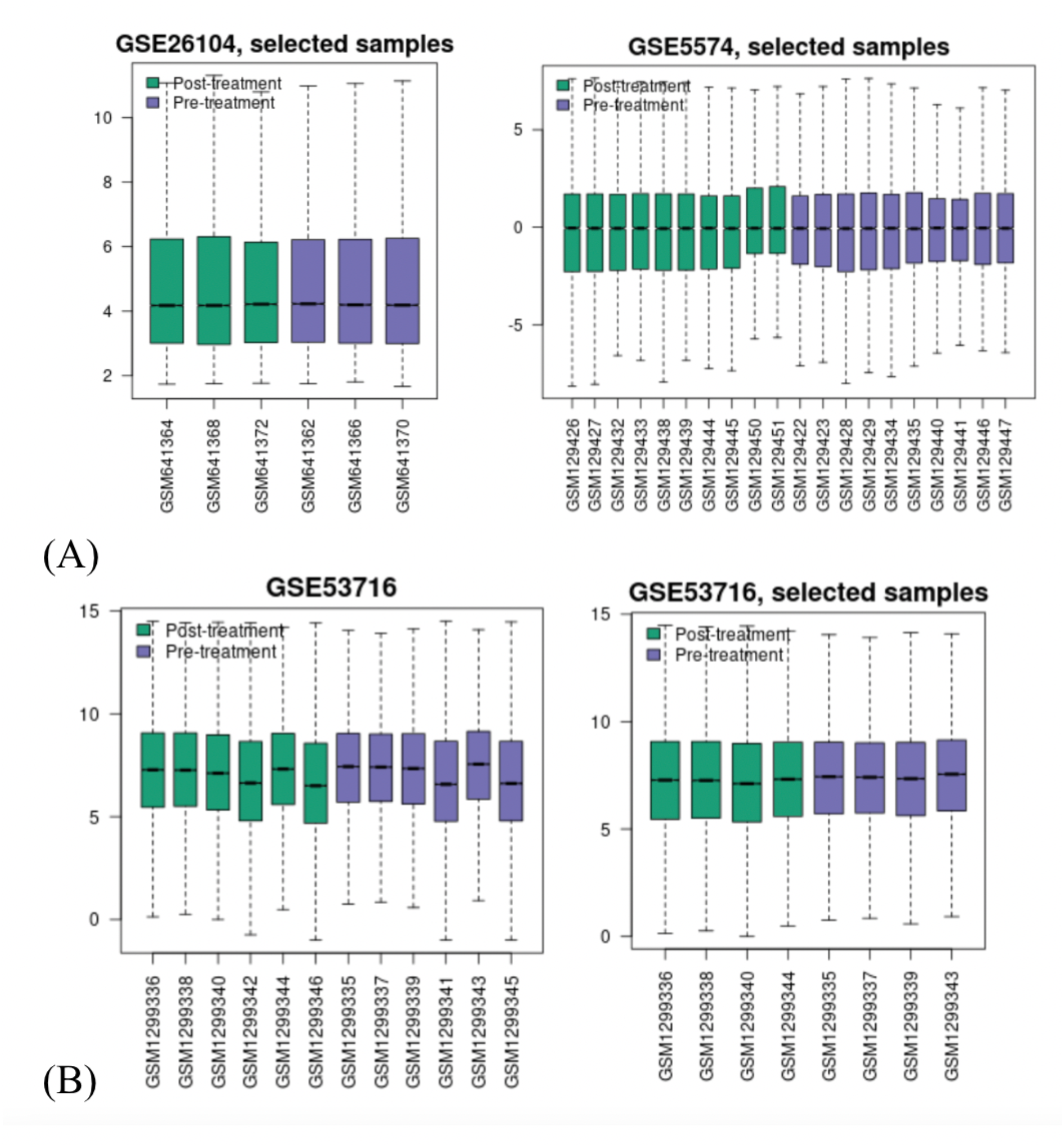
Gene expression value distribution. In figure (A), Gene expression value distribution for GSE26104 (left) and GSE5574 (right) and in (B) Gene expression value distribution for GSE53716 before (left) and after (right) data normalization by removing outliers manually. The vertical axis represents expression values of the genes (no unit). Each box plot represents the gene expression values of one patient sample denoted on the horizontal axis.

We considered p-value p < 0.05 and | log_2_FC (logFC) | ≥ 1 to be statistically significant for the DEGs, and a logFC ≥ 1, logFC ≤ −1 are considered to indicate up-regulated and down-regulated DEGs, respectively. We processed the lists of DEGs in Excel, where we removed DEGs with p-values of greater than 0.05 and the remaining DEGs are sorted into two categories: overexpression and underexpression. Next, only genes with | logFC | ≥ 1 are kept (Supplementary).

### Functional Enrichment of Gene Sets

The initial ontology of gene of the DEGs was annotated (p < 0.05) using the online bioinformatics tool DAVID (v2022q4, https://david.ncifcrf.gov/ accessed on 25 February 2023). DAVID provides exploratory visualization tools that promote discovery through functional classification, biochemical pathway maps, and conserved protein domain architectures from large lists of genes [14]. The human genome was selected as the background parameter, and official gene symbol was selected as identifier. The KEGG pathway enrichment analyses of the DEGs are cross-checked using PANTHER database provided by Gene Ontology (GO) (http://geneontology.org/ accessed on 25 February), which is a large database that stores information on biological pathways, components, functions, and the involved genes [15]. GO study is a frequently used approach for the functional studies of large-scale transcription or genomic data. Accordingly, GO’s tool PANTHER allows us to process a provided list of genes and calculates which biological entities those genes are overrepresented in [16].

### Construction of Protein - Protein Interaction (PPI) Network

The online database STRING (v11.0, http://www.string-db.org/ accessed on 23 February 2023) was used to construct the PPI network of the proteins encoded by DEGs. The STRING is an online repository with 24,584,628 proteins from 5090 organisms to predict the relationship between genes [17].

### Selection of Central Hub Proteins from PPI Network

A tab separated value file containing the gene graph in text form was exported and imported into Cytoscape. Cytoscape is a desktop-based tool that can perform more powerful analyses and functions on networks than STRING. The Network Analyzer tool in Cytoscape was used to calculate metrics of the gene network such as node degree and clustering coefficient [18]. Proteins that have a node degree at the top 10% of the sample and with a degree of ≥ 10 are considered hub proteins. The node degree is the number of genes connected to a certain gene. Additionally, the Molecular Complex Detection MCODE was used within Cytoscape to discover significant modules in the protein interaction network. MCODE is a graph clustering algorithm that is able to identify “densely connected regions” in networks [19].

## Result

### DEGs Identification

We base our study on our proposed methodology and the cut-off criteria (| log_2_FC | ≥ 1 and p-value p < 0.05). In GSE5574, 110 genes passed the criteria. In GSE26104, 331 genes passed the criteria, and in GSE53716, 3088 genes passed the criteria. The genes from each of the three datasets are inputted into JVenn (http://jvenn.toulouse.inra.fr/app/example.html accessed on 25 February 2023) to generate a three-way Venn diagram and identify overlaps (Figure 2) [20]. The majority of RNA was present in only one study (2337 out of 2436, around 96%), despite the fact that the processed data had p-values of < 0.05. 99 genes are found in two out of three datasets and 28 genes are found in all three databases, and interestingly all of which are overexpressed in post-treatment samples. The 28 overlapping RNAs are selected for further analysis and their logFC values are retrieved from each dataset and shown in a heat map that reveals the recurrently upregulated RNAs, including IFI44L, SERPING1, USP18, HESX1, and IFI44 (Figure 3).We will further analyse the significant DEGs after identifying the hub proteins.

**Figure 2.**
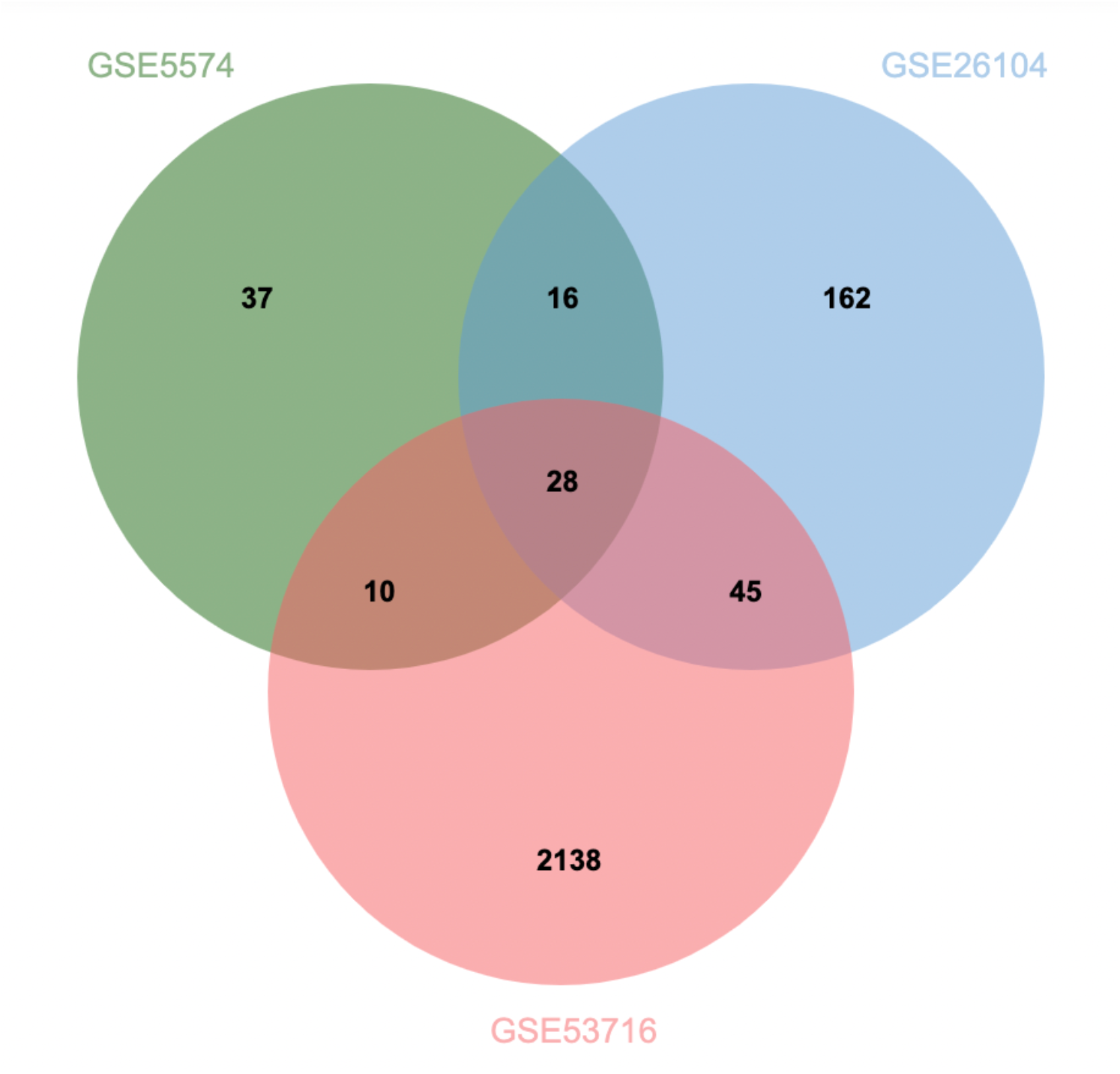
Three-way Venn diagram displaying overlapping RNA between the three chosen datasets.

**Figure 3.**
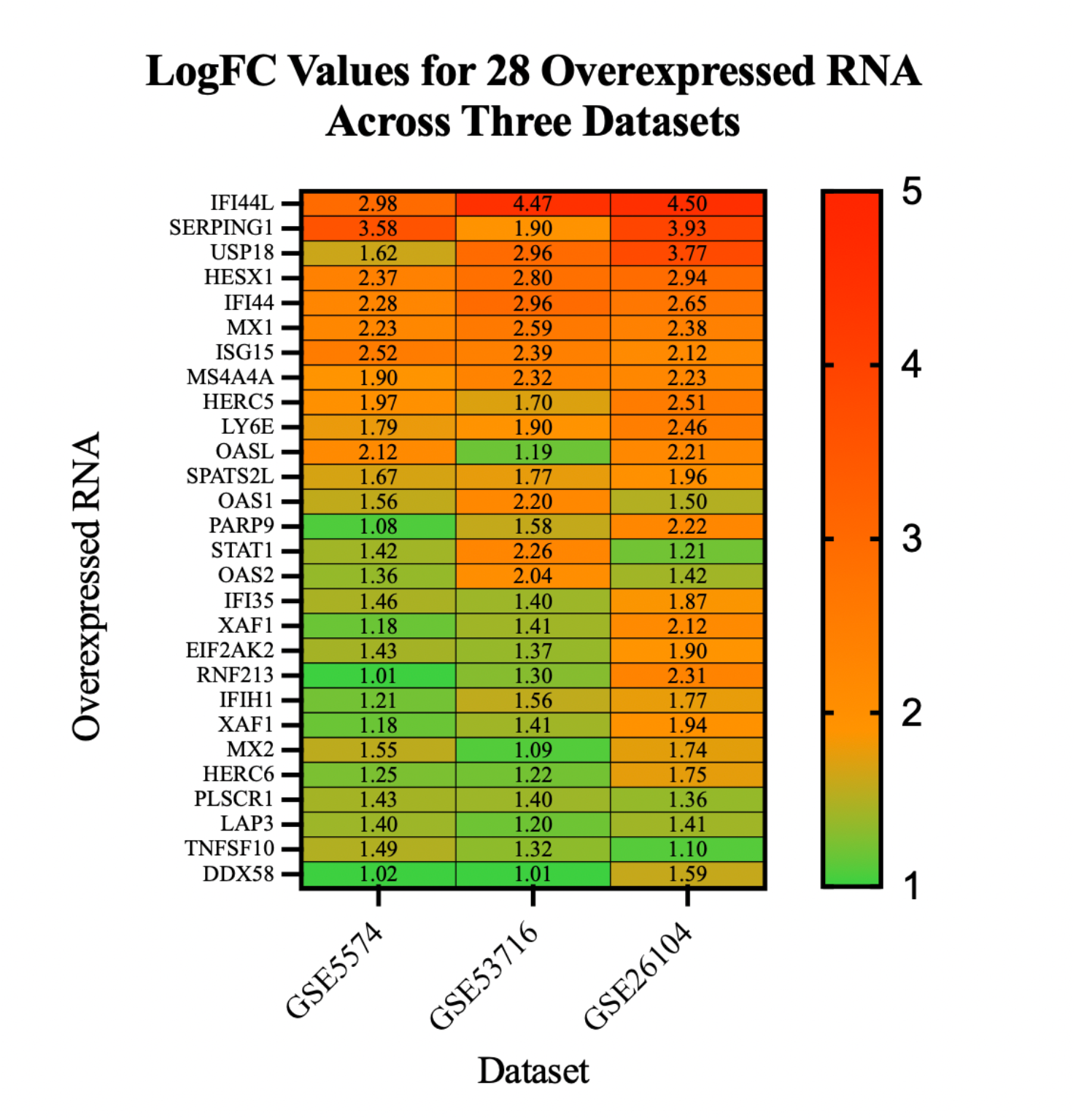
Heatmap of logFC values of the 28 over-expressed RNAs in post-treatment samples. Colour code red to green with red indicating a greater magnitude of expression (more positive logFC value). The RNAs are plotted in order of greatest average positive value based on average logFC in the three datasets for visualization. All logFC values displayed on the heatmap are statistically significant (p < 0.05).

### Functional Analysis of DEGs

DAVID analysis revealed several significantly enriched GO biological processes and molecular functions. The top 10 enriched biological processes and functions are displayed for the significantly enriched DEGs are depicted in Figure 4a. This is cross-analysed with PANTHER database, where the top 10 enrichment analysis outcomes are screened for the 28 DEGs in Figure 4b. The more enriched the pathway, the more biological significant it is.

**Figure 4.**
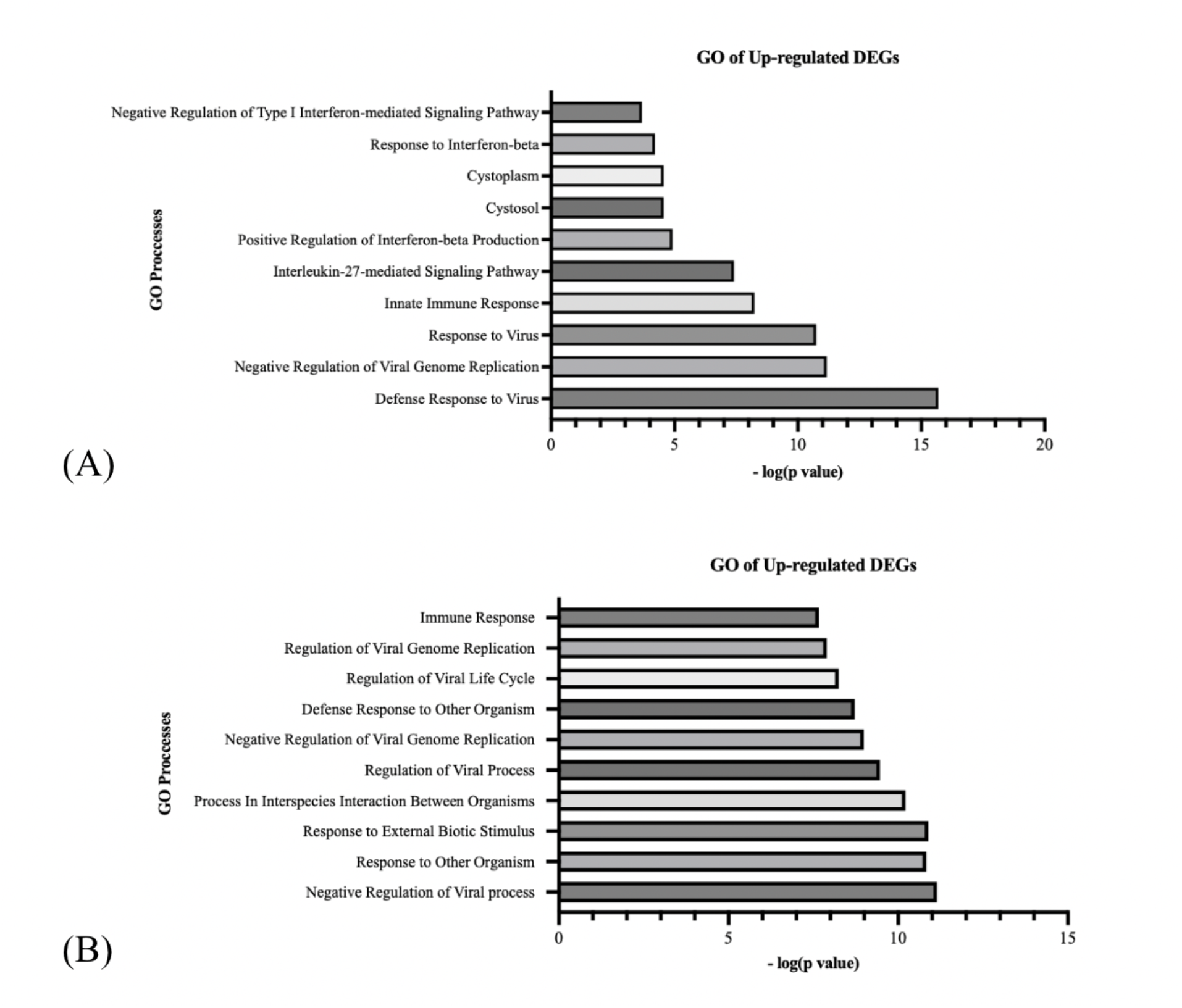
Functional analyses of the upregulated DEGs in post-treatment samples with (A) DAVID (B) PANTHER. GO processes include that of biological process, cellular component, and molecular function.

### PPI Network Construction

To evaluate the PPIs between the DEGs, we used the STRING tool to identify the PPI networks for upregulated DEGs. The 28 DEGs are submitted to the STRING tool. Solitary genes are hidden to improve clarity. The minimum confidence score for whether a protein interaction existed was set to 0.400, but we set the score to 0.700 (high confidence) to ensure higher reliability and clarity of our results, creating Figure 5. There are 148 interactions (edges) between 28 genes (nodes) with an average node degree of 10.6. The expected number of interactions was only 1 so the network had significantly more interactions than would be expected (p < 1.0E-16).

**Figure 5.**
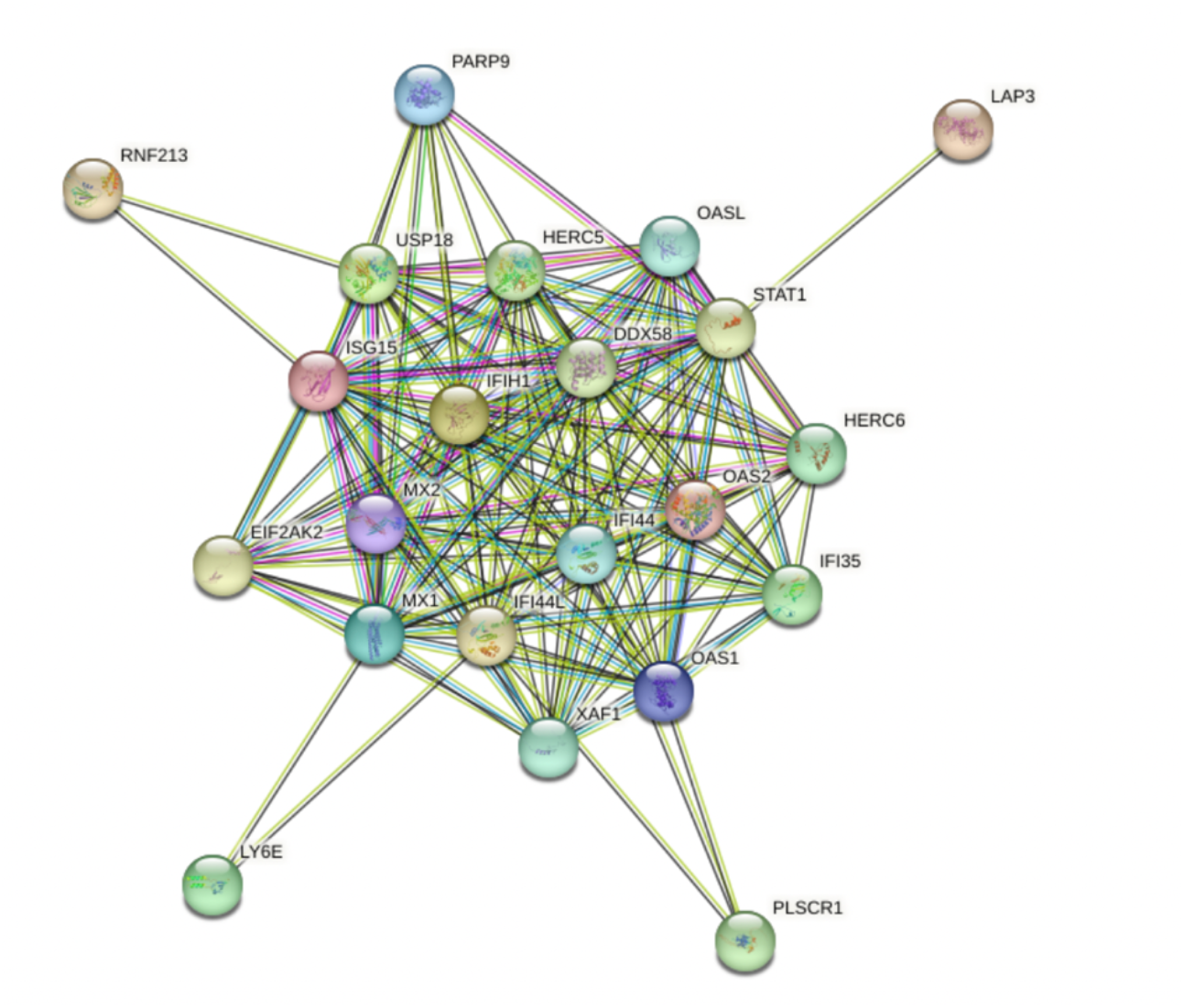
PPI network generated by STRING. Disconnected nodes are hidden and minimum interaction confidence is set to 0.700 for clarity. Each circle represents one gene and its protein product. Lines between circles are “edges” and represent interactions.

Expectedly, 12 out of 28 DEGs are involved in “innate immune” with a false discovery rate (FDR) of 4.28e-12 and 10 out of 28 DEGs are involved in “Interferon alpha/beta signalling” with a FDR of 5.63e-15.

Interestingly, 14 out of the 28 highly-expressed DEGs in post-treatment groups are linked to “antiviral defence” with a FDR of 1.28e-20 and 6 out of 28 DEGs are involved in “Epstein-Barr virus infection’ with a FDR of 2.67e-05. Since the involvement of viral infection in MS has been of great interest, we colour-coded the DEGs involved in “antiviral defence” and “Epstein-Barr virus infection” for further analysis. Figure 6a shows that the proteins involved in the aforementioned biological processes are interconnected. We also altered the confidence level to 0.900 (highest confidence) and about the same DEGs are interconnected (Figure 6b).

**Figure 6.**
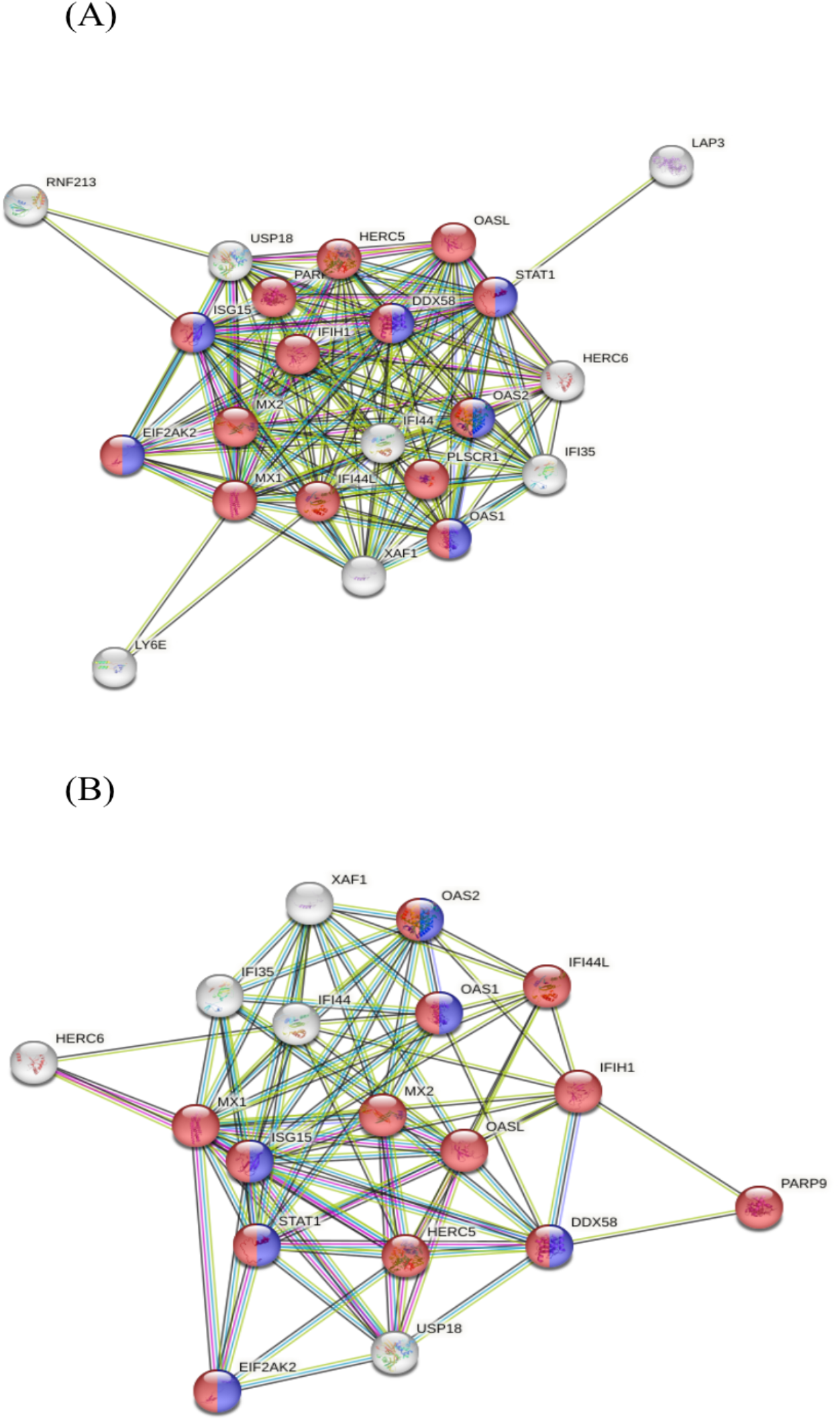
PPI network generated by STRING. Disconnected nodes are hidden and minimum interaction confidence is set to 0.900. Each circle represents one gene and its protein product.

Lines between circles are “edges” and represent interactions. Red nodes represent involvement of protein in “antiviral defence”, blue nodes represent involvement of protein in “Epstein-Barr virus infection”. In (A), the confidence level is set at 0.700. In (B), the confidence level is set at 0.900.

### Modules and Hub Proteins Identification

The resulting PPI network from STRING was exported as a “.txt” file and imported as a.csv file into Cytoscape v3.9.1 software for visualization. We have generated a merged network of PPI consisting of 28 nodes and 217 edges. The clustering coefficient is 0.922 out of 1, which indicates a dense connection.

MCODE was used to detect modules (highly interconnected clusters of genes) in the Cytoscape network. One module was identified by the MCODE algorithm. The module is composed of 20 genes with 187 interactions between them. The DEGs in the module is shown in Figure 7. Functional enrichment analysis of the module with PANTHER revealed that 14 of 20 genes in the module are involved in “Defence Response to Virus” with a fold enrichment of 56.97, and 13 of 18 genes are involved in “Immune Response”. In particular, “interleukin-27-mediated signalling pathway”, “regulation of ribonuclease activity”, “regulation of type III interferon production”, “cellular response to exogenous dsRNA”, and “ISG15-protein conjugation” are enriched for >100 folds (not shown).

**Figure 7.**
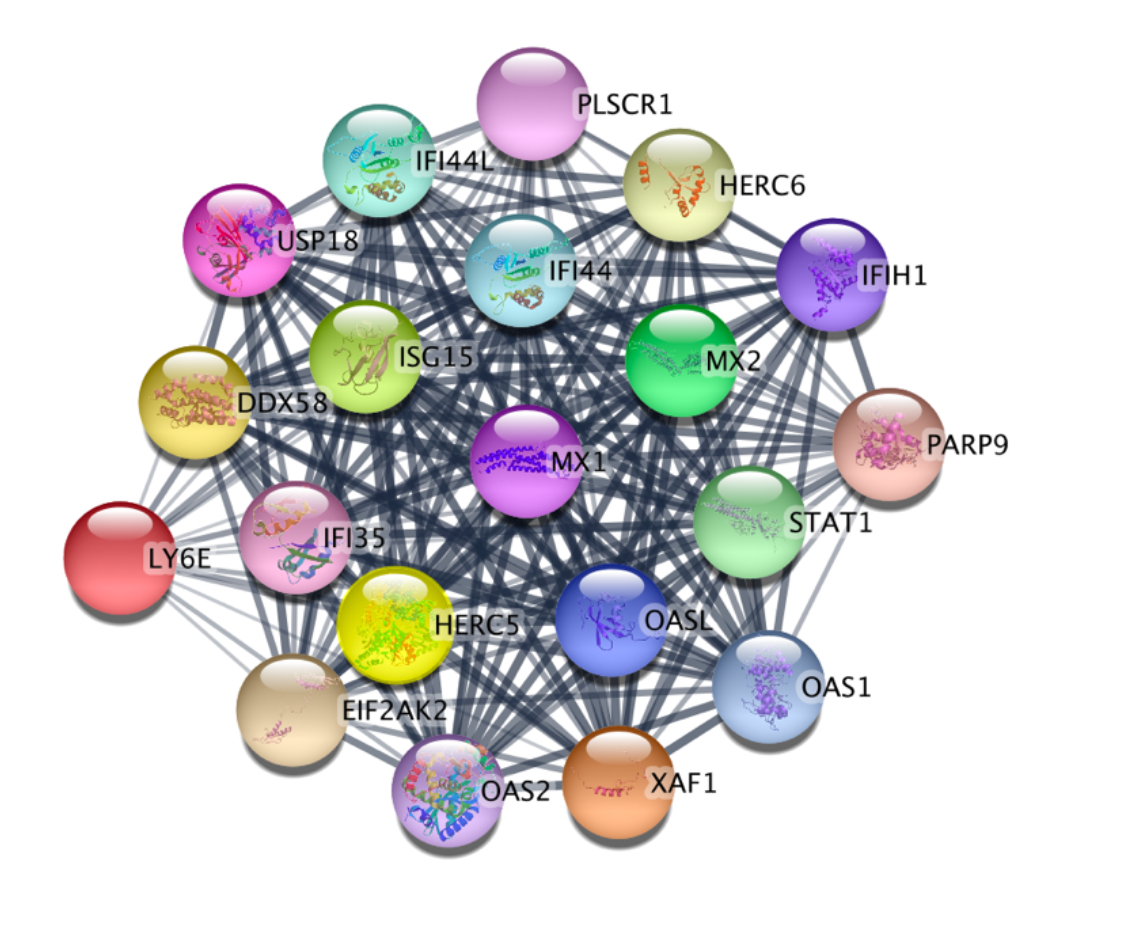
Module analysis of PPI network. The module has 20 nodes and 187 edges.

Next, the tool Network Analyzer in MCODE was used to select hub genes (minimum node degree 10 and top 10% of sample). Accordingly, 3 genes passed the hub gene cut-off criteria shown in Table 2. Interestingly, IF144L and IF144 are within the top five of the most enriched DEGs in the post-treatment samples (Figure 3).

**Table 2.** Cytoscape calculated node degrees for the genes in the network. The genes with node degrees of at least 10 and at the top 10% are presented: IFI144, IFI44L, and STAT1. The corresponding protein for each gene is also listed.

| Hub Gene | Protein | Node Degree |
| --- | --- | --- |
| IFI44 | Interferon-induced protein 44-like | 23 |
| IFI44L | Interferon-induced protein 44 | 22 |
| STAT1 | Signal transducer and activator of transcription 1-alpha/beta | 22 |

## Discussion

There are relatively few studies of gene expression during IFNβ therapy of MS patients [5]. Our analysis of three datasets on MS gene expression pre- and post-treatment yielded 99 DEGs that are found across a minimum of two studies, while 28 DEGs are found across all three datasets, which are all over-expressed in the post-treatment samples. These genes pass the criteria for p-value (p < 0.05) and fold change (| log_2_FC | ≥ 1) and are attractive for further research as they are biologically relevant.

From the pathway analysis with DAVID and PANTHER database, we have found that most of the pathways are mainly concentrated in “immune response”, “defence against virus”, and “regulation of viral genome replication” (Figure 4). This is supported by PPI network analysis showing high involvement of DEGs in immunity and defence against virus, especially where DEGs related to “antiviral defence” and “Epstein-Barre virus (EBV) infection” are interconnected in the PPI (Figure 5, 6). The involvement of the DEGs in EBV infection is of interest because recent study has found out there is a higher rate of EBV infection among people who developed MS than among controls [21]. Yet, this study does not show a causal relationship between the two, and further study is warranted to elucidate whether EBV infection increases risks of MS.

In module analysis with MCODE and PANTHER, we found out “interleukin-27-mediated signalling pathway”, “regulation of ribonuclease activity”, “regulation of type III interferon production”, “cellular response to exogenous dsRNA”, and “ISG15-protein conjugation” are enriched for >100 folds, suggesting the significance of the protein module in modulating these biological processes to affect the immunomodulary outcome of IFN-β (not shown).

In terms of immunomodulation, three biological pathways have been enriched for >100 folds. Firstly, “interleukin-27-mediated signalling pathway” was enriched for >100 folds. The role of interleukin-27 (IL-27) in regulating T cell responses that prevent immune hyperactivity has been extensively characterized and IL-27 has been investigated as a possible therapeutic for chronic inflammatory conditions with excessive T cell activation, which includes MS [22]. On the other hand, the anti-inflammatory capability of IL-27 can also be induced from other cell types in the central nervous system such as macrophages, microglia, and dendritic cells [23]. Overall, the finding suggests the MOA of interferon-β and suggests other possible therapies targeting IL-27 in alleviating MS symptoms. Secondly, “regulation of ribonuclease activity” was also enriched for >100 folds. Ribonucleases (RNases) are RNA-processing or - degrading enzymes that hydrolyse phosphodiester bonds within RNA molecules [24]. The contribution of RNases in inflammation modulation has been documented increasingly. For example, studies report that RNase T2 can act as an alarmin, which is an alarm-like molecule that acts on the innate immune system to send “dangerous” signals (such as bacterial infection, tissue damage, etc) [25]. Research has also shown that high expression of RNase protects against immune activation and inflammation such as in systemic lupus erythematosus, which is also an autoimmune disease [26-27]. Yet, the research on the role of RNases in MS is limited and more research is warranted. Thirdly, “regulation of type III interferon production” was also enriched for >100 folds. Type III interferons (IFNL) are antimicrobial cytokines that play key roles in immune host defence at endothelial and epithelial barriers. IFNLs signal via their heterodimeric receptor, comprised of two subunits, IFNLR1 and interleukin 10Rβ. Recent work has shown that IFNL might play an important role in the pathogenesis of immune-related disease including MS. In the experimental autoimmune encephalomyelitis model of MS, IFNLR1-/- animals demonstrated improved clinical disease course and decreased spinal cord axonal injury compared with WT animals [28]. Therefore, this might suggest a MOA for IFNβ’s immunomodulatory effect due to increased regulation of IFNL to alleviate MS symptoms and accordingly, more research is warranted to develop drugs targeting IFNL to treat autoimmune disease.

In terms of defence against virus, two biological pathways have been enriched for >100 folds. Firstly, “cellular response to exogenous dsRNA” was enriched for >100 folds. Since the presence of exogenous double-stranded RNA (dsRNA) is usually indicative of a viral infection, we can infer that IFNβ play a role in modulating the cell’s response to viral infection [29]. Secondly, “ISG15-protein conjugation” was also enriched for >100 folds. ISG15 is a member of the ubiquitin family and directly inhibits viral replication and modulates immune response. ISG15 can be covalently conjugated onto target proteins via an enzymatic cascade, yet the fate of these modified proteins is still largely unknown [30].

In our PPI analysis, we identified 3 hub genes based on node degree value from the merged PPI analysis. This included IFI44L, IFI44, and STAT1. It is to note that OAS1, OAS2, OASL IFIH1, IFI35, MX1, MX2, USP18, ISG15, XAF1, EIF2AK2, PLSCR1 had 20 interactions with other genes, meaning that they might be of interest as well. For example, the gene MX1 encodes for Mycovirus A (MxA), a protein with antiviral activity, and has been proven to be a sensitive measure of IFNβ bioactivity in MS [9].

For our 3 hub genes, IFI44L is a gene of 26 kb, larger than the 14 kb of IFI44, but both genes encode similar-sized proteins translated from a transcript produced from nine exons. IFI44 is made up of 444 amino acids, whereas IFI44L has 452 residues; the two proteins share 45% amino acid identity. Overexpression of IFI44 has been shown to restrict Bunyamwera virus and HIV-1 infection in vitro [31]. Yet this was first identified in the context of hepatitis C virus infection. IFI44L has been shown to have a moderate impact on hepatitis C virus infection. Interestingly, IFI44L expression has also been associated with several autoimmune disorders, cancer, and humoral responses to vaccination. These seemingly disparate contexts suggest that IFI44L is a biomarker of IFNβ responses independent of the type of stimulus. On the other hand if we consider the enriched pathways for antiviral defence from the functional analysis, this could also suggest a relationship between MS and viral infections that is yet to be unpacked with future research. STAT1 is also an identified hub gene which encodes for a protein with 750 residues. Expectedly, it plays a role as a signal transducer and transcription activator to mediate cellular responses to IFNs, cytokines, and other growth factors [32].

Limitations in this study are that the overexpressed genes in MS patients are not compared to that of healthy control within the 12-month span due to a lack of data from the GEO databases (including GSE53716 and GSE5574). Only GSE26104 has compared the gene expression before and after treatment of IFNβ for MS patients and healthy controls, where USP18 was found the only biomarker found to be differentially expressed between MS patients and controls, suggesting that USP18 may play a role in the pathogenesis of MS [9]. In our study, USP18 is one of the 28 DEGs present in all 3 databases with a node degree of 20, suggesting that proteins that are not identified as hub protein cannot be overlooked. Due to the “scale-free” nature of protein interaction network, there is no consensus in the literature on the degree threshold that defines a hub protein. Given the largely ad hoc definition of hub proteins, it is possible that many special properties attributed to hubs may be simply a consequence of the definition used [33]. From our results, although further studies are needed, it is tempting to speculate that overexpression of IF144L and IF144 in MS patients in post-treatment compared to pre-treatment samples has implications for the therapeutic effect of IFNβ. Lastly, the research databases did not include the responsiveness of patients to the IFNβ-1a therapy after 12 months due to “the clinical decline in our non-responder occurred prior to collecting the chronic treatment specimen” in GSE5574 [10]. Although it has been estimated that determination of response requires lengthy clinical follow-up of up to 2 years, further research is warranted to identify whether these overexpressed genes after IFNβ treatment are linked to clinical responsiveness and non-responsiveness of MS patients towards IFNβ to figure out biomarkers for the responsiveness towards IFNβ treatment [6].

## Conclusion

The present study identified 28 RNAs that are overexpressed in post-treatment MS patients in all three datasets. Functional enrichment analysis of the DEGs revealed several overrepresented pathways relevant to immune defence and antiviral activity. 3 target genes are hub genes in the PPI network and possess significant prognostic value. More research on the correlation of gene expression and responsiveness towards IFNβ treatment and the correlation between MS and viral infection are warranted. Although our results do not prove that the changes in these genes in MS patients and in IFN-β-dosed MS patients are anything but coincidental, this abnormal pathway may be a window into the etiology or immune pathogenesis of MS.

## Data Availability

This study was using pre-existing gene expression data available freely from the NCBI Gene Expression Omnibus, PANTHER Database, DAVID Database and STRING Database.

## Supplemental

For the pages in excel, “All” represents all the genes that pass the cut-off criteria, which is p- value p < 0.05 and a | log_2_FC | ≥ 1. “Overexpressed” are the genes with log_2_FC ≥ 1 while “Underexpressed” are the genes with log_2_FC ≤ 1.

### GSE26104 Excel

https://docs.google.com/spreadsheets/d/1aGlJoMPEuxwXKt9XpXMVpfCerIIkJUNhHrSMWBBsr1E/edit?usp=sharing

### GSE5574 Excel

https://docs.google.com/spreadsheets/d/15NKGwozlsbXVrrNW9quyXnHFWPjY9Mp43rcU3_0axTw/edit?usp=sharing

### GSE53716 Excel

https://docs.google.com/spreadsheets/d/1g9_AWuuyDDiHUvLW2qFLrV4Fn5ghJvb_bOHpGrhitXs/edit#gid=1364008547

## Notes

### Competing Interest Statement

The authors have declared no competing interest.

### Funding Statement

This study did not receive any funding

### Author Declarations

This study was using pre-existing gene expression data available freely from the NCBI Gene Expression Omnibus.

### Summary of Updates

The abstract is slightly miswritten (one extra differentially expressed genes is documented) and the Discussion is being refined slightly

